# Title and abstract screening for systematic reviews with Jev, a System One model: comparison with generative large language models

**DOI:** 10.64898/2026.09.25.26364021

**Authors:** Kentaro Matsui, Yoshikazu Takaesu

**Affiliations:** Department of Sleep Medicine, National Center Hospital, National Center of Neurology and Psychiatry, Tokyo, Japan; Department of Sleep-Wake Disorders, National Institute of Mental Health, National Center of Neurology and Psychiatry, Tokyo, Japan; Department of Neuropsychiatry, Graduate School of Medicine, University of the Ryukyus, Okinawa, Japan

**Keywords:** systematic reviews, title and abstract screening, machine learning, large language models, probability thresholds, workload reduction

## Abstract

Large language models (LLMs) screen titles and abstracts without review-specific training, but generating screening decisions as text takes processing time and incurs API charges. We evaluated Jev, a non-generative model returning classification probabilities, on 4527 records from two systematic reviews of bipolar disorder treatments. We used human reviewers’ decisions to retain records for full-text assessment as the reference standard. In the primary analysis, we asked Jev yes-or-no questions (“Noul”) about whether to retain each record. Records were retained when their retention probability was at least 50%, a cutoff specified before the final evaluation runs. We also evaluated lower retention thresholds. For comparison, we gave Jev explicit include/exclude options (“Choice”) and used the same prompts with GPT-5 mini and GPT-6 Astra; both LLMs used default reasoning. In the primary analysis at 50%, sensitivity and specificity were 99.6% and 98.9% for light therapy and 93.2% and 93.5% for adjunctive pharmacotherapy. Lower cutoffs selected on these data retained all reference-positive records. Compared with assessing every record manually, these cutoffs reduced the number requiring human assessment by 96.9% for light therapy and 61.3% for adjunctive pharmacotherapy. For light therapy and adjunctive pharmacotherapy, sensitivity was 92.3% and 90.0% with Jev using explicit options at 50%, 87.2% and 90.0% with GPT-5 mini, and 92.3% and 80.8% with GPT-6 Astra. Costs for screening all 4527 records once were US$0.20 for Jev using explicit options, US$2.59 for GPT-5 mini, and US$33.45 for GPT-6 Astra. Jev’s probabilities could provide graded information for reviewers assessing titles and abstracts.

**Highlights:** *What is already known:* Generative LLMs can screen titles and abstracts using natural-language eligibility criteria, but generating each decision takes time and incurs API charges.

*What is new:* Jev screened records without generating text or requiring training on records labeled for each review. In two reviews, screening cost substantially less than with the tested LLM configurations.

*Potential impact for Research Synthesis Methods readers:* Rather than providing only an include-or-exclude decision, Jev indicates how strongly the model favors retaining each record. Reviewers could consider this information alongside the title, abstract, and eligibility criteria.

## 1. Introduction

Clinical practice guidelines depend on systematic reviews, which take a long time to produce,^1^ and title and abstract screening accounts for much of the workload. With earlier machine-learning classifiers, records were ranked by a relevance score, but training required labeled records from each review.^2,3^ Large language models (LLMs) can screen titles and abstracts using eligibility criteria written in natural language, without training on records labeled for the individual review. Title and abstract screening is now conducted with LLMs, and recent evidence syntheses reported high sensitivity.^4,5^ Although routine use of LLMs has been judged premature,^6^ combining judgments from several models or pairing an LLM with a human reviewer performed well,^7,8^ and implementation has already extended to national clinical guideline production.^9^

Screening, however, is a classification task performed through text generation with generative LLMs. Outputs are decisions rather than scores^10–12^ and LLM confidence is often poorly calibrated.^13–15^ Errors also occur: eligible studies were excluded under some screening conditions with GPT-5, a previous frontier model from OpenAI.^16^ In addition, generating each screening decision and any intermediate reasoning token by token takes processing time and incurs API charges.^17–19^ The cost of screening a fixed set of records can be lowered by sending multiple records to the LLM in each request (batching). However, increasing the number of records per request has been reported to reduce sensitivity or produce invalid outputs in some models.^20^

Jev, a recently introduced non-generative model developed by TypeSafe AI and described by its developer as a System One model, processes natural-language input and returns probabilities for specified answers without generating text token by token.^21,22^ According to TypeSafe AI, Jev evaluates multiple questions about the same input in parallel, so asking additional questions has little effect on the time needed to return a response.^22^ For screening, several records can be placed in the shared input, with a separate question asking whether to retain each record. This arrangement could reduce the time and cost of screening a fixed set of records. Like an LLM, Jev does not require training on records labeled for each review. It also provides a score for each record, a feature of earlier classifiers trained for individual reviews,^2,3^ which may convey more information than a binary include-or-exclude decision.

Although Jev has shown task-dependent probability calibration when used to judge generated responses,^23^ its screening performance and the correspondence between its probabilities and human retention decisions remain untested in systematic reviews. We evaluated Jev on two datasets from our previous study of title and abstract screening^10^ and compared Jev with two generative LLMs using the same prompts. We examined sensitivity, the number of records requiring human review, and probability calibration against human retention decisions. We also assessed whether performance was maintained as the number of records per request increased, and measured latency, processing time, and cost.

## 2. Methods

### 2.1. Study design, datasets, and reference standard

We analyzed two datasets previously used to evaluate GPT-3.5 and GPT-4^10^: 1381 records from a systematic review of bright light therapy for bipolar disorder (hereafter, light therapy)^24^ and 3146 from a systematic review of adjunctive pharmacotherapy for bipolar depression.^25^ Eligibility criteria are described in the source reviews,^24,25^ and human screening procedures in our previous study.^10^ The original records had been screened in duplicate by pairs of human evaluators. Their decisions formed the basis of the reference standard; records retained for full-text assessment are referred to as human-retained records. In our previous study, records retained by human reviewers but excluded by GPT were reassessed. When exclusion was judged appropriate, the reference label was changed to excluded. We incorporated 13 adjudicated exclusions and resolved three additional labels: a related open-label report, a discrepancy between source sheets, and a missing consensus label. Reference labels were then fixed before the final evaluations.^10^ The final reference standard comprised 26 human-retained records for light therapy and 87 for adjunctive pharmacotherapy.

### 2.2. Jev and prompts

A Jev request contains the content to be evaluated, termed the state, and one or more questions. Each question consists of instructions and a question type. The questions are evaluated against the same state independently and in parallel, and each receives an answer in the format specified by its type, without text generation. Figure 1 summarizes the screening approaches and study design. We used two question types. A “Noul” question asks for a yes-or-no judgment and returns a single value from 0 to 1: the probability of yes. A “Choice” question presents a set of options and returns a probability for each option; these probabilities sum to 1.^21–23^ We accessed jev-1.13.0 through the TypeSafe API on September 18, 2026 (UTC).

**Figure 1.**
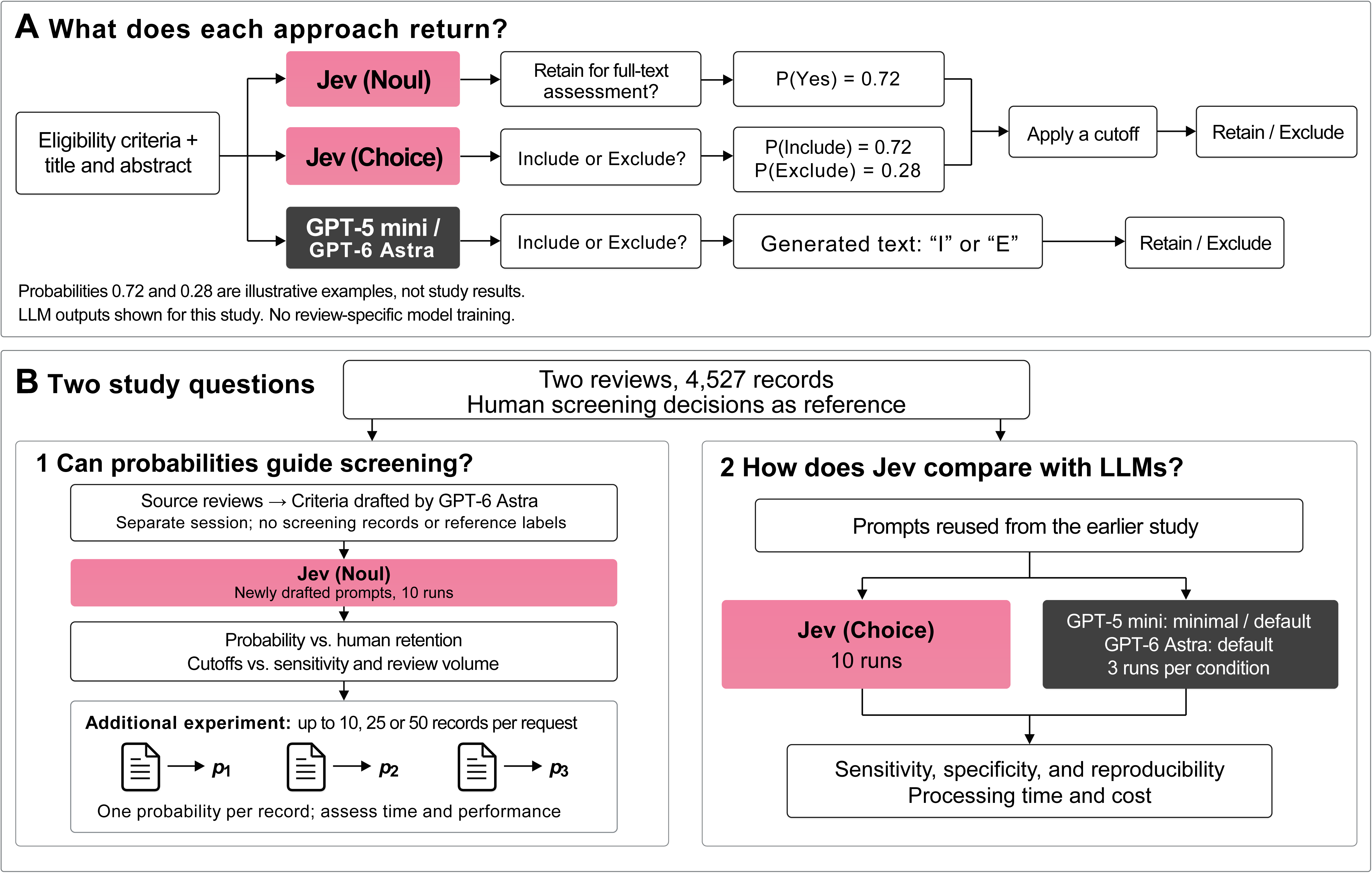
Screening approaches and study design. Panel A illustrates the inputs and outputs of the screening approaches. The output from Noul is the probability of a yes answer; outputs from Choice include probabilities for the specified options. For Jev, the probability of retention was compared with a cutoff. Outputs from Choice also include the most probable option and a confidence value, neither of which was used in this analysis. LLM outputs were I or E under the study settings. Panel B shows the two analyses: evaluation of Jev probabilities against human screening decisions using Noul and newly drafted prompts, and comparison with generative LLMs using Choice and prompts adapted from the earlier study. Additional Noul experiments evaluated multiple records per request. Human retention refers to retention for full-text assessment, not final inclusion. Noul and Choice were not compared using identical prompts. LLM denotes large language model.

The primary analysis used Noul questions.^22^ We used GPT-6 Astra (OpenAI) as an agent in a separate Codex session to draft screening criteria from the two source reviews,^24,25^ without giving GPT-6 Astra access to screening records, reference labels, earlier prompts, or screening results. The criteria specified that records with uncertain eligibility should be retained and clearly incompatible records excluded. Each request’s state contained the criteria and one or more records, with an identifier for each record. One Noul question per record asked whether that record should be retained for full-text assessment. We treated the returned value as the probability of retention.

The secondary analysis used Choice questions.^22^ We adapted the prompts from our previous study^10^ to assess study design, population, intervention, and comparator together, with include and exclude as the options. Each request’s state contained one record, and the instructions contained the criteria and query. We used the probability assigned to include as the probability of retention. The complete prompts are provided in Figure S1.

### 2.3. Screening runs

A run consisted of screening all 4527 records across both datasets once. As the focus of this study, Jev was run 10 times under each condition to characterize run-to-run variability. The primary analysis used one record per request, and we repeated it with up to 10, 25, or 50 records per request. The concurrency limit was 50 requests in all conditions. Single-record runs used a fixed input order. For requests containing multiple records, each dataset was reshuffled before every run using fixed seeds, supplied with the code. The input budget was approximately 32,000 tokens per request.^26^ To stay within this budget, we grouped records using a 90,000-character budget for their combined title and abstract text, including the title and abstract labels. Transient failures were retried automatically. All returned identifiers and probabilities were checked programmatically for completeness and valid format.

### 2.4. Comparison with generative LLMs

We accessed GPT-5 mini (gpt-5-mini-2025-08-07) and GPT-6 Astra (gpt-6-astra) through the OpenAI API on September 19, 2026 (UTC). Each request contained one record, followed by the criteria, query, and response options of the secondary analysis and an instruction to answer with the single letter I or E.^10^ We ran GPT-5 mini with the lowest accepted reasoning setting (minimal) and the default setting (reasoning effort unspecified), and GPT-6 Astra with the default setting. Previous studies set the temperature to 0 to reduce variability in outputs,^10^ but the API did not accept a temperature setting for either model. We therefore ran each condition three times, with up to 50 concurrent requests. For all models, request latency covered dispatch to the parsed response, excluding pacing and retry waits. Run processing time included pacing, rate-limit waits, and retries. Costs were calculated from token usage in saved successful responses at the study list prices, including LLM reasoning tokens as output tokens. Setup calls and unobserved usage from failed requests were excluded. Time and token usage were counted once per request when records were bundled.

### 2.5. Statistical analysis

We calculated sensitivity and specificity for each dataset and run, using human screening decisions as the reference. We also calculated the percentage reduction in records requiring human assessment compared with reviewing all records. A 50% probability cutoff was specified before the final evaluation runs: Jev retained records with a probability at or above this cutoff. We evaluated other cutoffs in 5-percentage-point increments. For each dataset, we also identified the highest cutoff, to the nearest percentage point, at which Jev retained every human-retained record in all 10 runs.

We grouped records by Jev’s probability of retention and calculated the percentage in each group that human reviewers had retained for full-text assessment. To use the same probability intervals in the primary and secondary analyses, we first calculated each record’s mean probability across runs in each analysis. We used these means to count records in 10-percentage-point intervals, then merged adjacent intervals until each contained at least 20 records in both analyses. After fixing these boundaries, we assigned records to intervals using their probabilities in each individual run. We summarized the percentages retained by human reviewers as means and ranges across runs.

We measured variation between runs as the percentage of all 4527 records for which the retain-or-exclude decision differed between runs. Jev was run 10 times and each LLM condition three times. To compare variation over the same number of runs, we considered all 120 possible combinations of three Jev runs. For each combination, we calculated the percentage of records whose decisions were not identical across the three runs, then averaged these 120 percentages. We also calculated the standard deviation of each record’s probability across the 10 Jev runs, separately for each condition defined by the number of records per request.

## 3. Results

### 3.1. Screening performance in the primary analysis

After full-text assessment, six reports of six trials were included in the original light-therapy review, and nine reports of five trials in the original adjunctive-pharmacotherapy review. Jev Noul retained all of these reports in all 10 runs at the prespecified 50% cutoff. In the primary analysis at the prespecified 50% cutoff, mean sensitivity and specificity were 99.6% and 98.9% for light therapy and 93.2% and 93.5% for adjunctive pharmacotherapy. For light therapy, 49% was the highest cutoff that retained all human-retained records in every run. At this cutoff, mean specificity was 98.7% and the number of records requiring human assessment was reduced by 96.9%. For adjunctive pharmacotherapy, the corresponding cutoff was 20%, with mean specificity of 63.1% and a reduction in review volume of 61.3% (Table 1).

**Table 1.** Screening with Jev Noul and newly drafted prompts at different probability cutoffs (primary analysis).

| Retention cutoff | Sensitivity,<br>% | Specificity,<br>% | Records retained | 100% sensitivity,<br>runs |
| --- | --- | --- | --- | --- |
| **Light therapy (1381 records; 26 retained by the reference standard)** |  |  |  |  |
| ≥10% | 100.0 | 72.9 (72.8 to 73.1) | 392.7 (391 to 395) | 10/10 |
| ≥20% | 100.0 | 88.9 (88.5 to 89.5) | 176.2 (168 to 182) | 10/10 |
| ≥30% | 100.0 | 95.5 (95.3 to 95.7) | 87.1 (84 to 90) | 10/10 |
| ≥50%<sup>a</sup> | 99.6 (96.2 to 100.0) | 98.9 (98.7 to 99.0) | 41.4 (40 to 43) | 9/10 |
| ≥49%<sup>b</sup> | 100.0 | 98.7 (98.6 to 98.9) | 43.1 (41 to 45) | 10/10 |
| **Adjunctive pharmacotherapy (3146 records; 87 retained by the reference standard)** |  |  |  |  |
| ≥10% | 100.0 | 30.9 (30.6 to 31.0) | 2201.4 (2198 to 2210) | 10/10 |
| ≥20% | 100.0 | 63.1 (62.6 to 63.4) | 1217.0 (1208 to 1232) | 10/10 |
| ≥30% | 98.9 | 79.7 (79.4 to 80.0) | 708.5 (699 to 717) | 0/10 |
| ≥50%<sup>a</sup> | 93.2 (93.1 to 94.3) | 93.5 (93.4 to 93.7) | 279.6 (273 to 284) | 0/10 |
| ≥20%<sup>b</sup> | 100.0 | 63.1 (62.6 to 63.4) | 1217.0 (1208 to 1232) | 10/10 |

Note: Values are means (range) across 10 runs; a single value indicates no variation at the reported precision. Records meeting the cutoff are retained for full-text assessment. The cutoff applies to the probability of retention. <sup>a</sup> Cutoff fixed before the final evaluation runs. <sup>b</sup> Highest cutoff retaining every human-retained record in all 10 runs, determined from these data.

Across the 10 primary-analysis runs, seven distinct human-retained records had a retention probability below 50% at least once: one from the light-therapy review and six from the adjunctive-pharmacotherapy review. A post hoc assessment of their titles and abstracts identified explicit incompatibilities with the primary-analysis criteria in three records; eligibility remained uncertain in the other four, warranting further assessment under the instruction to retain uncertain records (Table S1).

### 3.2. Human retention according to model-assigned probability

In both datasets and both analyses, no record assigned a probability below 10% was human-retained in any run. Among light-therapy records with probabilities of 60 to 100%, the mean percentage of human-retained records was 83.4% in the primary analysis and 93.6% in the secondary analysis. For adjunctive pharmacotherapy, the percentage of human-retained records was at most 4.9% in any interval below 60%. The percentage increased across the four intervals from 60% to 100%, reaching 85.9% in the primary analysis and 69.2% in the secondary analysis in the highest interval, 90 to 100% (Figure 2).

**Figure 2.**
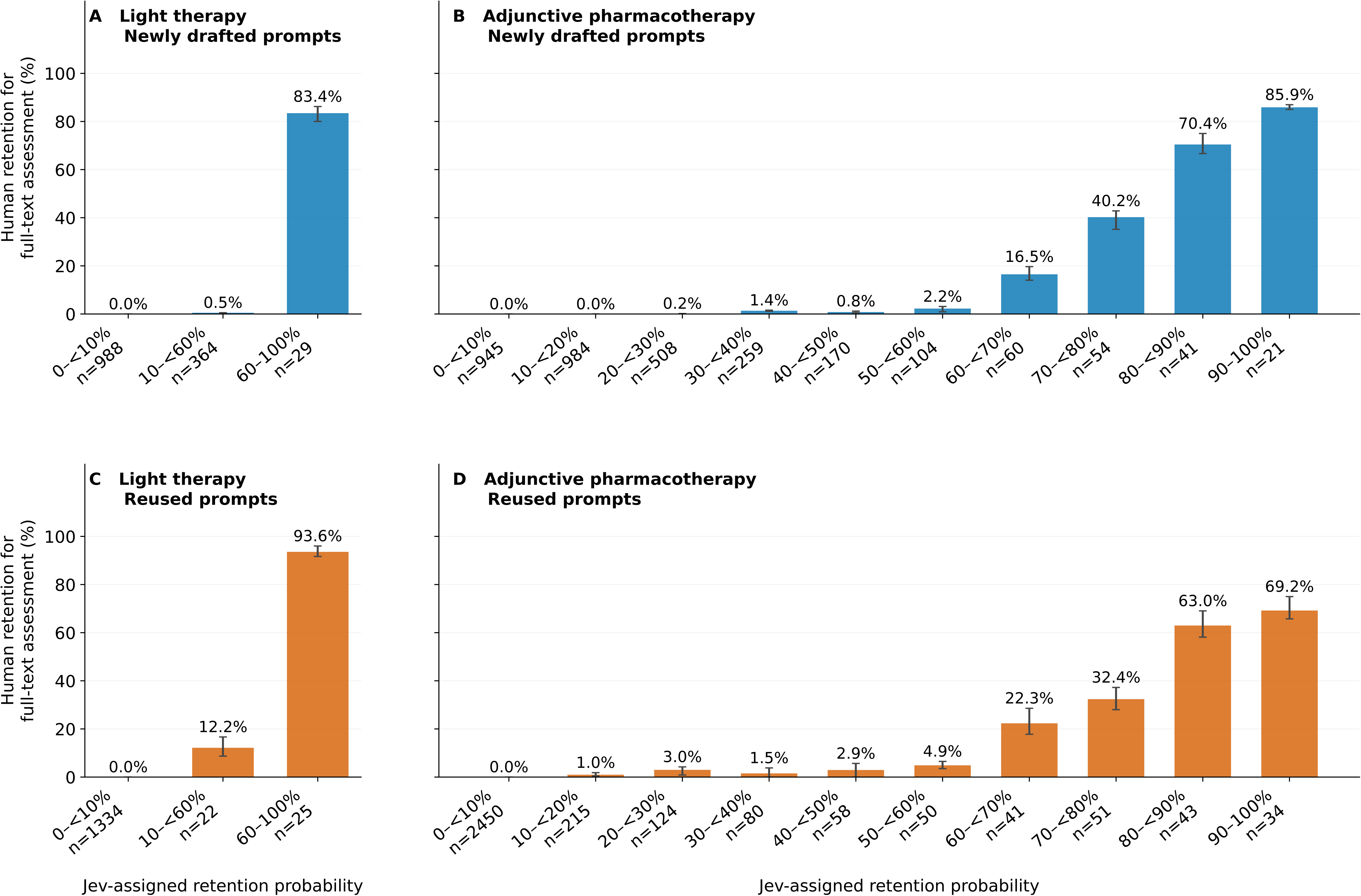
Human retention for full-text assessment according to Jev-assigned probability. Panels A and B show the results with the newly drafted prompts (primary analysis), and Panels C and D those with the prompts adapted from the earlier GPT-based study (secondary analysis). Bars indicate means across 10 runs, and error bars the range; n denotes the mean number of records per run. Intervals are shared between analyses within each dataset and displayed at equal spacing.

### 3.3. Comparison with generative LLMs

Jev Choice retained all reports included in the two systematic reviews in all 10 runs at the 50% cutoff. In the secondary analysis with Jev Choice at the 50% cutoff, sensitivity and specificity were 92.3% and 99.7% for light therapy and 90.0% and 95.4% for adjunctive pharmacotherapy. The highest cutoff retaining all human-retained light-therapy records in every run was 22%, with specificity of 99.2% and a reduction in review volume of 97.3%. For adjunctive pharmacotherapy, the corresponding cutoff was 14%, with specificity of 83.4% and a reduction in review volume of 81.1% (Table 2).

**Table 2.** Screening with Jev Choice and generative LLMs using the same adapted prompts (secondary analysis).

| Model and retention cutoff | Sensitivity,<br>% | Specificity,<br>% | Records retained | Human-retained<br>records excluded |
| --- | --- | --- | --- | --- |
| **Light therapy (1381 records; 26 retained by the reference standard)** |  |  |  |  |
| Jev Choice, $\geq 10\%$ | 100.0 | 98.4 (98.4 to 98.5) | 47.1 (46 to 48) | 0.0 |
| Jev Choice, $\geq 20\%$ | 100.0 | 99.1 (99.0 to 99.1) | 38.3 (38 to 39) | 0.0 |
| Jev Choice, $\geq 30\%$ | 93.5 (92.3 to 100.0) | 99.4 (99.3 to 99.5) | 32.3 (31 to 34) | 1.7 (0 to 2) |
| Jev Choice, $\geq 50\%$<sup>a</sup> | 92.3 | 99.7 (99.6 to 99.9) | 28.0 (26 to 29) | 2.0 |
| Jev Choice, $\geq 22\%$<sup>b</sup> | 100.0 | 99.2 (99.1 to 99.2) | 37.3 (37 to 38) | 0.0 |
| GPT-5 mini, minimal reasoning | 73.1 (65.4 to 76.9) | 99.9 | 20.3 (18 to 22) | 7.0 (6 to 9) |
| GPT-5 mini, default reasoning | 87.2 (84.6 to 88.5) | 99.5 (99.4 to 99.6) | 30.0 (29 to 31) | 3.3 (3 to 4) |
| GPT-6 Astra, default reasoning | 92.3 | 98.1 (98.1 to 98.2) | 49.3 (49 to 50) | 2.0 |
| **Adjunctive pharmacotherapy (3146 records; 87 retained by the reference standard)** |  |  |  |  |
| Jev Choice, $\geq 10\%$ | 100.0 | 80.1 (79.9 to 80.4) | 695.6 (687 to 702) | 0.0 |
| Jev Choice, $\geq 20\%$ | 97.6 (95.4 to 98.9) | 87.1 (86.8 to 87.3) | 480.6 (474 to 489) | 2.1 (1 to 4) |
| Jev Choice, $\geq 30\%$ | 93.3 (92.0 to 94.3) | 91.0 (90.8 to 91.1) | 356.4 (352 to 361) | 5.8 (5 to 7) |
| Jev Choice, $\geq 50\%$<sup>a</sup> | 90.0 (89.7 to 90.8) | 95.4 (95.3 to 95.7) | 218.0 (211 to 221) | 8.7 (8 to 9) |
| Jev Choice, $\geq 14\%$<sup>b</sup> | 100.0 | 83.4 (83.1 to 83.7) | 593.3 (587 to 604) | 0.0 |
| GPT-5 mini, minimal reasoning | 70.9 (70.1 to 71.3) | 93.4 (93.2 to 93.7) | 262.7 (254 to 270) | 25.3 (25 to 26) |
| GPT-5 mini, default reasoning | 90.0 (89.7 to 90.8) | 93.2 (93.0 to 93.4) | 287.3 (280 to 291) | 8.7 (8 to 9) |
| GPT-6 Astra, default reasoning | 80.8 (80.5 to 81.6) | 94.4 (94.3 to 94.5) | 240.3 (237 to 245) | 16.7 (16 to 17) |

Note: Values are means (range) across three runs for GPT-5 mini and GPT-6 Astra and 10 runs for Jev; a single value indicates no variation at the reported precision. We used prompts adapted from the earlier GPT-based study for all models; we used Choice questions for Jev. The Jev cutoff applies to the probability of retention. With Jev, all human-retained records were retained in 10/10 runs at cutoffs of 10%, 20%, and 22% for light therapy and 10% and 14% for adjunctive pharmacotherapy; this occurred in 1/10 runs at 30% for light therapy and in 0/10 runs at the other displayed cutoffs. Mean processing time and cost per run were 137 seconds and US\$0.88 for GPT-5 mini with minimal reasoning, 270 seconds and US\$2.59 with default reasoning, 161 seconds and US\$33.45 for GPT-6 Astra, and 251 seconds and US\$0.20 for Jev. <sup>a</sup> Cutoff fixed before the final evaluation runs. <sup>b</sup> Highest cutoff retaining every human-retained record in all 10 runs, determined from these data. Abbreviations: LLM, large language model.

GPT-5 mini with minimal reasoning excluded one report included in the light-therapy review in all three runs and two or three reports included in the adjunctive-pharmacotherapy review per run. With the secondary-analysis prompts and minimal reasoning, mean sensitivity and specificity for GPT-5 mini were 73.1% and 99.9% for light therapy and 70.9% and 93.4% for adjunctive pharmacotherapy.

GPT-5 mini with default reasoning excluded the same light-therapy report in one of three runs and retained all nine reports included in the adjunctive-pharmacotherapy review in every run. With default reasoning, GPT-5 mini had sensitivity of 87.2% and specificity of 99.5% for light therapy; for adjunctive pharmacotherapy, sensitivity was 90.0% and specificity was 93.2%.

GPT-6 Astra with default reasoning retained all included reports from both reviews in all three runs. For GPT-6 Astra with default reasoning, sensitivity and specificity were 92.3% and 98.1% for light therapy and 80.8% and 94.4% for adjunctive pharmacotherapy.

Although we could not set the temperature to 0 for either LLM, all responses consisted of the single letter I (retain for full-text assessment) or E (exclude), as requested. Mean output tokens per record were 10 for GPT-5 mini with minimal reasoning, 200 for GPT-5 mini with default reasoning, and 9 for GPT-6 Astra.

Of the 113 records retained by human reviewers for full-text assessment across both datasets, Jev Choice excluded 10 or 11 per run at the 50% cutoff. GPT-5 mini excluded 31 to 35 with minimal reasoning and 11 to 13 with default reasoning; GPT-6 Astra excluded 18 or 19. We also examined Jev Choice’s probabilities for records excluded by the LLMs. With default reasoning, the two LLMs excluded 24 distinct human-retained records at least once across their three runs. For 10 of these 24 records, Jev Choice’s mean retention probability across 10 runs was below 50%.

We assessed run-to-run variability in retain-or-exclude decisions across repeated screenings of the same 4527 records. At the 50% cutoff, decisions differed between runs for 1.4% of records with Jev Choice and 2.6% with Jev Noul across all 10 runs. For the GPT models, each assessed over three runs, the corresponding percentages were 3.1% for GPT-5 mini under minimal reasoning, 4.6% under default reasoning, and 0.5% for GPT-6 Astra. To compare variability over three runs, we averaged the percentage of records with differing decisions across all 120 combinations of three Jev runs. At the 50% cutoff, these averages were 0.7% for Jev Choice and 1.4% for Jev Noul.Median request latency for Jev was 256 ms in the primary analysis and 258 ms in the secondary analysis. For GPT-5 mini, median latency was 1273 ms with minimal reasoning and 2332 ms with default reasoning; for GPT-6 Astra, it was 1511 ms. With one record per request, Jev requests were spaced to remain within the API request limit. This pacing determined the total processing time: a mean of 249 seconds per run in the primary analysis and 251 seconds in the secondary analysis. Mean processing time was 137 seconds for GPT-5 mini with minimal reasoning, 270 seconds with default reasoning, and 161 seconds for GPT-6 Astra. No request to any model received a rate-limit response.

Jev cost US$0.22 per run in the primary analysis and US$0.20 in the secondary analysis. Costs per run were US$0.88 for GPT-5 mini with minimal reasoning, US$2.59 with default reasoning, and US$33.45 for GPT-6 Astra.

### 3.4. Multiple records per request

At the 50% cutoff, Jev Noul retained all reports included in the two systematic reviews after full-text assessment. This was the case in all 10 runs with up to 10, 25, or 50 records per request. With limits of 10, 25, and 50 records per request, requests contained a mean of 10.0, 24.9, and 43.7 records, respectively. Every run completed without failed or invalid responses. Mean processing time for all 4527 records was 25.7 seconds with up to 10 records per request, 15.7 seconds with up to 25, and 15.3 seconds with up to 50, compared with 249 seconds with one record per request. Costs ranged from US$0.10 to US$0.11 per run across the multiple-record conditions. Median request latency was 450 ms with up to 10 records per request, 736 ms with up to 25, and 1270 ms with up to 50.

At the 50% cutoff, light-therapy sensitivity was 100.0% in every run across these conditions. Specificity ranged from 97.0 to 97.2%, compared with 98.9% with one record per request. For adjunctive pharmacotherapy, sensitivity was 91.7%, 90.5%, and 89.7% with up to 10, 25, and 50 records per request, respectively, compared with 93.2% with one record. Specificity ranged from 92.1 to 93.4%, compared with 93.5% with one record (Figure 3). At the highest cutoffs retaining all human-retained records, adjunctive-pharmacotherapy specificity was 51.1%, 57.7%, and 53.6% with up to 10, 25, and 50 records per request, respectively, compared with 63.1% with one record. The mean standard deviation of each record’s probability across runs was 0.009 with one record per request and 0.031 to 0.034 with multiple records per request.

**Figure 3.**
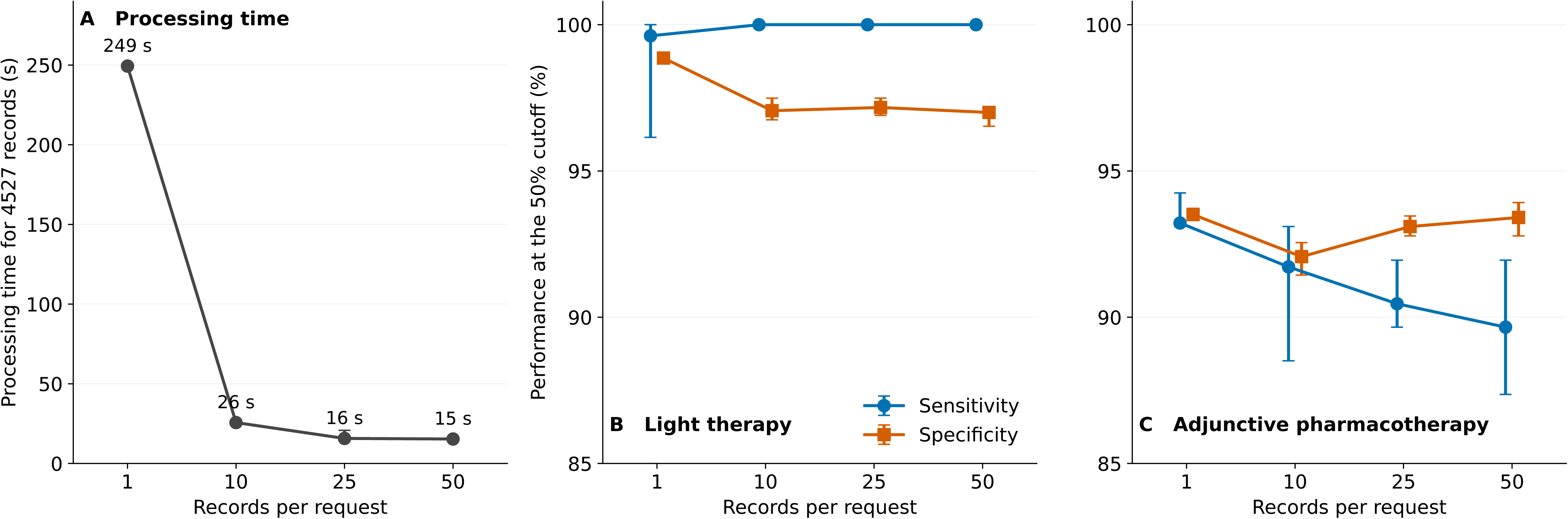
Screening of multiple records per request with Jev and the newly drafted prompts. Panel A shows the processing time per run for all 4527 records. Panels B and C show sensitivity and specificity at the 50% cutoff. Points indicate means across 10 runs, and error bars the range.

## 4. Discussion

In these reviews, Jev screened titles and abstracts without generating text or training on records labeled for each review. Jev Choice used the same prompts as the LLMs; at the prespecified cutoff, its sensitivity and specificity were similar to or higher than those of the LLMs with default reasoning, and its cost was lower. Like earlier classifiers trained for individual reviews,^2,3^ Jev provides a graded score for each record. Jev can do so using eligibility criteria written in natural language.

Jev’s low screening cost warrants comparison with GPT-5 mini as another low-cost screening option. Previous studies have reported high but imperfect LLM screening sensitivity;^5,27,28^ our earlier GPT-based study also documented exclusions of human-retained records.^10^ Even with minimal reasoning, GPT-5 mini cost more than Jev Choice. Moreover, it excluded reports ultimately included in both source reviews. Default reasoning improved sensitivity and retained all ultimately included reports from the adjunctive-pharmacotherapy review, but the same light-therapy report missed in all three runs with minimal reasoning was still excluded in one of three runs. Although Jev also excluded some human-retained records at the prespecified 50% cutoff, it retained all reports ultimately included after full-text assessment in both source reviews across all tested conditions and runs. GPT-6 Astra also retained all of these reports in all three runs, but screening all 4527 records once cost approximately 170 times as much as with Jev Choice. Previous research has documented errors in human abstract screening, with an estimated miss rate of approximately 8–13% with a single reviewer, and even dual independent screening can miss around 0–3% of eligible studies.^29–31^ Given the inherent limitations of human performance, the low miss rate of Jev may already be at a level sufficient to replace a single human reviewer, although further research is needed because its performance may depend on domain knowledge that varies across fields. Another potential advantage is that, Jev’s probability output indicates how strongly the model favors retaining a record. While it does not establish eligibility or substitute for human judgment on final inclusion,^32^ reviewers could consider this graded information alongside the title, abstract, and eligibility criteria when assessing a record.

Processing time also matters when screening must be repeated after searches are updated.^33^ With one record per request, Jev returned individual responses faster than the LLMs. However, the total time to screen all records was similar because Jev requests were spaced to stay within its API limit. Sending multiple records per request reduced the total time to seconds. All Jev runs completed successfully, whereas invalid outputs have been reported for some generative LLMs with larger batches.^20^ Of note, compared with sending one record per request, sending multiple records lowered sensitivity for adjunctive pharmacotherapy at the fixed cutoff and increased variation in probabilities across runs. Given that screening with one record per request already completed within a practical time frame, sending multiple records per request may not be necessary when sensitivity is prioritized.

Several limitations affect the interpretation of these findings. First, both reviews concerned bipolar disorder, limiting generalizability to other clinical areas. The datasets also contained few human-retained records, and no independent external validation was performed. Second, the highest cutoffs retaining all human-retained records were identified from the evaluation data and are therefore optimistic; they would not be known in advance in a prospective review. Third, the LLM comparison covered only two models from one developer and the tested reasoning settings. In addition, screening sensitivity may vary with prompt wording,^27,34^ but we did not systematically evaluate this variation. Finally, all models were accessed through proprietary APIs on specific dates, and the results may change with later model updates.

## 5. Conclusions

Across the two systematic reviews evaluated, Jev achieved high screening sensitivity at the prespecified cutoff, retained all reports included after full-text assessment, and cost less than the tested LLM configurations. External validation is needed, as this study used only the datasets from our previous evaluation. System One models such as Jev are likely to play a central role in title and abstract screening for future systematic reviews. The emergence of additional models in this class could broaden the options for simplifying systematic review workflows and improving review quality.

## Acknowledgments

We used GPT-6 Astra (OpenAI) and Claude Fable 5.1 (Anthropic) to assist with prompt development, code drafting, analysis, and manuscript preparation. These tools were accessed through Codex or Claude Code. The screening models and their use are described in the Methods. The authors are responsible for the final content.

## Author contributions

Kentaro Matsui: Conceptualization, Methodology, Software, Formal analysis, Data curation, Writing - original draft, Funding acquisition. Yoshikazu Takaesu: Data curation, Supervision, Writing - review & editing.

## Funding

Japan Society for the Promotion of Science (JSPS) KAKENHI, grant 26K10503. The funder had no role in study design, data collection, analysis, or interpretation.

## Competing interests

The authors declare none.

## Ethics

This study analyzed bibliographic records and existing screening labels; no human participants or identifiable patient data were involved. Ethics approval and participant consent were not applicable.

## Data availability

Reference labels, model predictions, request-level resource measurements, prompts, model settings, and analysis code are available at https://github.com/matsuikentaro1/jev-title-abstract-screening. These materials support reanalysis of the saved model outputs. Original titles and abstracts include third-party database content, including Embase, and are not shared because of applicable licensing restrictions. Repeating model inference requires the original input texts and access to the relevant APIs.

## Notes

### Competing Interest Statement

The authors have declared no competing interest.

