## Supplementary Appendix (Figure S1 and Table S1) for "Title and abstract screening for systematic reviews with Jev, a System One model: comparison with generative large language models"

**Contents**

Figure S1. Screening prompts (page 2)

Table S1. Human-retained records excluded in at least one primary-analysis run (page 3)

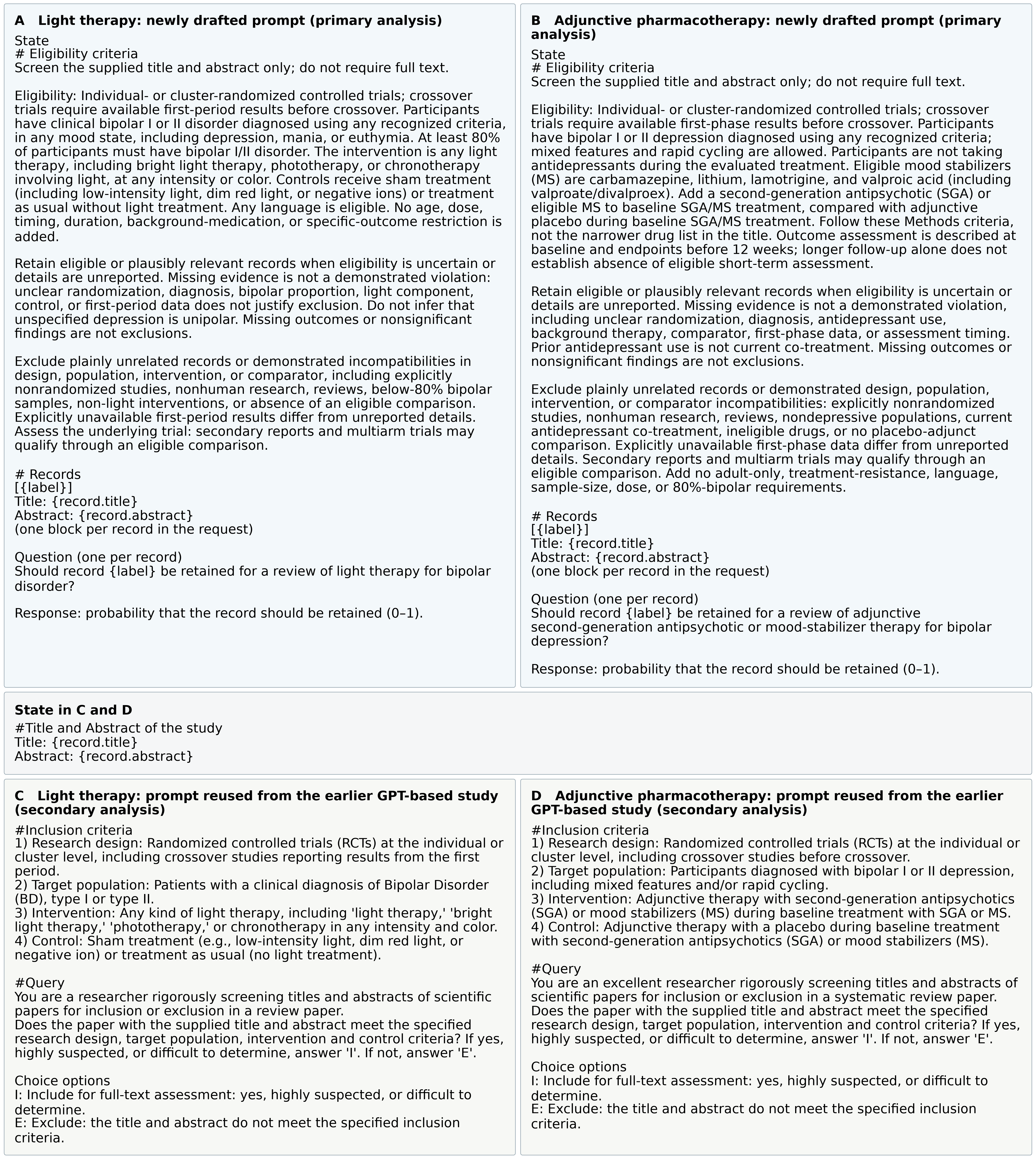

***Figure S1.*** *Screening prompts. Panels A and B show the prompts newly drafted from the eligibility criteria of each review (primary analysis), and Panels C and D the prompts adapted from the earlier GPT-based study (secondary analysis). Prompts for light therapy are on the left and those for adjunctive pharmacotherapy on the right. Braced fields are replaced with record text. GPT-5 mini and GPT-6 Astra received the record and the text of Panel C or D in one message, followed by the instruction to answer with the single letter I or E.*

***Table S1.*** *Human-retained records excluded by Jev Noul at the 50% cutoff in at least one primary-analysis run.*

| **Record and source** | **Retention, %  Excluded runs** | **Summary of the supplied title and abstract** | **Post hoc assessment against primary-analysis criteria** |
| --- | --- | --- | --- |
| 1. Morning light treatment hastens the antidepressant effect of citalopram: a placebo-controlled trial  Light therapy  [PMID: 12823078](https://pubmed.ncbi.nlm.nih.gov/12823078/) | 53.7 (49 to 60)  1/10 | Thirty inpatients receiving citalopram were randomized to morning green light or an inactive negative-ion generator, with outcomes assessed over two weeks. Of these patients, nine had bipolar disorder and 21 had major depressive disorder. | **Exclusion is supported by the population criterion.** Only 9/30 patients (30%) had bipolar disorder, below the 80% minimum specified in the primary-analysis criteria. |
| 2. Risperidone does not affect steady-state pharmacokinetics of divalproex sodium in patients with bipolar disorder  Adjunctive pharmacotherapy  [doi:10.2165/00003088-200443110-00004](https://doi.org/10.2165/00003088-200443110-00004) | 22.8 (20 to 26)  10/10 | Twenty-two patients with bipolar disorder in remission received divalproex and were randomized to adjunctive risperidone or placebo for two weeks. The study assessed valproate pharmacokinetics and the safety of combined treatment. | **Exclusion is supported by the population criterion.** Patients were in remission, whereas the primary-analysis criteria required bipolar depression. This reported mood state supports exclusion; the focus on pharmacokinetics alone would not be sufficient. |
| 3. Aripiprazole as adjunct to a mood stabilizer and citalopram in bipolar depression: A randomized placebo-controlled pilot study  Adjunctive pharmacotherapy  [doi:10.1002/hup.1096](https://doi.org/10.1002/hup.1096) | 33.6 (30 to 39)  10/10 | Twenty-three inpatients with bipolar depression receiving lithium or valproate were randomized to adjunctive aripiprazole or placebo for six weeks. All patients also received citalopram during this period. | **Exclusion is supported by the antidepressant criterion.** Both groups received citalopram, whereas the primary-analysis criteria excluded antidepressant use during the evaluated treatment. The lack of a significant treatment benefit would not itself justify exclusion. |
| 4. Quetiapine Extended Release (XR) in Bipolar Patients With Comorbid Generalized Anxiety Disorder (GAD)  Adjunctive pharmacotherapy  [CENTRAL: CN-02033813](https://www.cochranelibrary.com/central/doi/10.1002/central/CN-02033813/full) | 34.1 (30 to 37)  10/10 | An eight-week randomized, double-blind trial was planned to compare quetiapine XR with placebo in 120 adults with bipolar I or II disorder and generalized anxiety disorder. The description included monotherapy or adjunctive mood-stabilizer therapy, with weekly mood assessments. | **Full-text or detailed registry assessment is warranted.** Although the trial focused on anxiety, the description does not establish that patients with bipolar depression were absent. It also includes a potentially eligible adjunctive comparison; the unspecified mood state therefore calls for further assessment under the instruction to retain uncertain records. |
| 5. A pilot study of lithium carbonate plus divalproex sodium for the continuation and maintenance treatment of patients with bipolar I disorder  Adjunctive pharmacotherapy  [doi:10.4088/jcp.v58n0301](https://doi.org/10.4088/jcp.v58n0301) | 36.7 (33 to 39)  10/10 | Twelve patients with bipolar I disorder receiving lithium were randomized to divalproex or placebo. Follow-up continued for up to one year to assess relapse or recurrence and adverse effects during continuation and maintenance treatment; additional medication was permitted as needed. | **Further assessment is warranted.** The focus on maintenance treatment makes exclusion understandable, but the description does not establish a definite violation of the criteria. Mood state at entry, short-term assessments, and additional medication are not sufficiently described to support exclusion under the instruction to retain uncertain records. |
| 6. A double-blind, placebo-controlled study with quetiapine as adjunct therapy with lithium or divalproex in bipolar I patients with coexisting alcohol dependence  Adjunctive pharmacotherapy  [doi:10.1111/j.1530-0277.2010.01270.x](https://doi.org/10.1111/j.1530-0277.2010.01270.x) | 40.2 (37 to 44)  10/10 | In a 12-week randomized placebo-controlled trial, 362 patients with bipolar I disorder and alcohol dependence received quetiapine or placebo added to lithium or divalproex. Heavy drinking was the primary outcome, and secondary measures included global illness severity. | **Further assessment is warranted.** The adjunctive intervention and comparator are potentially eligible, but whether patients had bipolar depression is unclear. The focus on alcohol use does not resolve this uncertainty, and the 12-week endpoint does not establish that earlier assessments were absent. |
| 7. Valproate Efficacy in Cocaine-Bipolar Comorbidity  Adjunctive pharmacotherapy  [CENTRAL: CN-01512077](https://www.cochranelibrary.com/central/doi/10.1002/central/CN-01512077/full) | 46.3 (43 to 50)  9/10 | The trial description compared adjunctive valproate with adjunctive placebo in patients with bipolar disorder and cocaine dependence. Both groups were to receive usual treatment. Although cocaine use was the main focus, the planned analyses included depressed versus manic/mixed subgroups and assessments of mood symptoms. | **Further assessment is warranted.** A depressed subgroup was specified in the planned analyses, but details of randomization and background treatment were insufficient. Because neither the focus on cocaine use nor these missing details establish a violation of the criteria, the record should be retained for further assessment. |

All seven records were retained for full-text assessment by the reference standard but received a retention probability below 50% in at least one of 10 primary-analysis runs with one record per request. Retention probabilities are shown as means (minimum to maximum) across 10 runs. Human retention refers to full-text assessment, not final inclusion in the source review. Source links identify the reports or database records. The summaries are based on the titles and abstracts used for screening.
